# Features of mobile apps for diabetic kidney disease self-management: a scoping review

**DOI:** 10.1101/2025.08.20.25334066

**Authors:** Hasmawati Yahya, Nani Draman, Azidah Abdul Kadir, Najib Majdi Yaacob

## Abstract

**Background:** Diabetic kidney disease (DKD) is a chronic complication of diabetes mellitus (DM). DKD and chronic kidney disease (CKD) are both long-term conditions requiring ongoing patient care. Individuals living with DKD or CKD can benefit from mobile apps that support effective self-management. However, limited evidence is available about what mobile apps features are commonly included for DKD.

**Objective:** This scoping review aimed to identify the features of mobile applications on self-management for individuals with DKD, DM or CKD.

**Methods:** The review followed the Joanna Briggs Institute (JBI) methodology for scoping reviews and adhered to PRISMA-ScR reporting guidelines. Five databases (PubMed, Scopus, SAGE Journals, ScienceDirect, and Web of Science) were searched from inception to February 26, 2025. Studies were included if they reported on mobile apps supporting self-management in adults with DKD, DM or CKD. DM and CKD apps were included due to similar self-management needs such as blood sugar or blood pressure tracking that are also relevant to people with DKD. Data were extracted on study characteristics, app features, use of technology, integration with care teams, and reported outcomes.

**Results:** Out of 3521 records identified, eleven studies met the inclusion criteria. Five studies focused on CKD, two on DKD, and four on diabetes. Across the eleven mobile apps reviewed, four core domains of self-management were identified: self-care monitoring (91%), educational components (64%), patient support and motivation (100%), and performance incentives (18%). Four apps employed wearable devices and incorporated supportive devices such as Bluetooth glucometers. However, only two apps included real-time communication features with providers integration with healthcare teams and gamification strategies.

**Conclusions:** Mobile apps targeting DKD frequently incorporate monitoring, education, and motivational features. However, consistent integration with healthcare providers and incentive-based engagement strategies remains limited. Future app development should emphasise personalised feedback, clinical integration, and sustained engagement mechanisms to enhance usability and impact.

## Introduction

Diabetes and chronic kidney disease (CKD) are two significant global health challenges that often coexist and interact. Diabetic kidney disease (DKD), a condition that emerges when diabetes leads to progressive kidney damage, is one of the most serious long-term complications of diabetes. It significantly increases the risk of kidney failure, cardiovascular disease, and premature death. Managing these conditions requires patients to take on various daily tasks, such as monitoring blood glucose or blood pressure, adhering to medications, making dietary changes, and engaging in healthy lifestyle behaviours. These self-management responsibilities can be overwhelming for many individuals, especially without structured support [1,2].

In recent years, mobile applications (apps) have gained attention as practical and scalable tools for self-management of chronic diseases like diabetes and CKD. These apps offer a variety of features that can help patients better understand their condition, track health metrics, receive medication or appointment reminders, and access educational content on diet, exercise, and symptoms. Because mobile phones are widely accessible and integrated into many people’s daily lives, mobile apps have become a convenient and potentially impactful way to enhance disease self-care [3].

Self-management refers to an individual’s ability to manage the symptoms, treatment, lifestyle changes, and psychosocial consequences of living with a chronic condition. It emphasizes active participation in care, including making informed decisions, adhering to treatment plans, and engaging in behaviors that promote health and well-being [4,5].In the context of diabetic kidney disease and chronic kidney disease effective self-management is linked to improved clinical outcomes, greater self-efficacy, and reduced healthcare utilization [6,7].

Despite the growing number of mobile apps developed for diabetes or CKD individually, relatively few are designed specifically for people with DKD. However, because DKD is closely linked to both diabetes and CKD, mobile apps built for either condition often include overlapping features such as blood glucose and blood pressure monitoring, medication reminders, educational content, and provider communication tools that are also relevant for DKD management [3,8]. Including mobile apps targeting diabetes, CKD, and DKD in this review enables a more comprehensive assessment of how digital health tools support self-management across these interrelated conditions [11–21].

Therefore, this scoping review aims to explore and map the key features of mobile applications developed to support self-management for people living with diabetes, CKD, or DKD. Looking across all three conditions helps provide a complete picture of how mobile technology supports important health behaviours like monitoring, education, motivation, and treatment adherence. By understanding what’s already available and how these features are used, this review hopes to highlight areas where mobile app development is doing well and where more tailored, patient-centred solutions may still be needed [2,3].

## Material and methods

This scoping review was conducted in accordance with the Joanna Briggs Institute (JBI) methodology for scoping reviews and reported following the Preferred Reporting Items for Systematic Reviews and Meta-Analyses extension for Scoping Reviews (PRISMA-ScR) checklist. The research questions in this scoping review were developed using the Joanna Briggs Institute’s Population–Concept–Context (PCC) framework to ensure alignment with scoping review methodology [S1 Table]. The population of interest includes individuals diagnosed with both diabetes and chronic kidney disease (CKD) or diabetic kidney disease (DKD). The concept explored is self-management, which refers to patients’ active participation in managing their health conditions. The context focuses on the use of mobile applications that support these self-management activities. The PCC elements guiding this review are summarized in Table 1.

**Table 1.** PCC Framework for the scoping review research questions.

| PCC Element | Keywords/Terms |
| --- | --- |
| Population | Diabetes AND (Chronic Kidney Disease OR Diabetic Kidney Disease) |
| Concept | Self-management |
| Context | Mobile applications (mobile apps) |

### Protocol and registration

No protocol was registered for this scoping review. However, the methodology followed the Joanna Briggs Institute (JBI) guidelines for conducting scoping reviews and adhered to the PRISMA-ScR (Preferred Reporting Items for Systematic Reviews and Meta-Analyses extension for Scoping Reviews) reporting checklist.

### Inclusion criteria

Studies was included if they describe or evaluate mobile applications (apps) designed to support self-management among individuals with diabetic kidney disease (DKD) or diabetes or chronic kidney disease. Articles must report on at least one self-management-related feature integrated into a mobile app, such as health monitoring, patient education, motivational tools, or performance feedback. Only studies published in English were included. There were no publication year, study design, or geographic location restrictions.

More specifically, the inclusion criteria were:

i. Population: Adults diagnosed with type 2 diabetes and chronic kidney disease (CKD) or diabetic kidney disease (DKD). Studies focusing on patients with both conditions, or where DKD is clearly defined, will be included.
ii. Concept: The main focus is self-management interventions delivered via mobile applications. This includes features such as self-care monitoring, education, motivation, support, and performance incentives.
iii. Context: Studies conducted in any healthcare or community setting, with mobile applications (not limited by platform) used to support self-management among DKD patients. No geographic restrictions exist, as long as the study includes relevant app-based interventions.
iv. Types of Studies: This scoping review included interventional studies (both randomised and non-randomised), observational studies (such as cohort and cross-sectional designs), and studies describing the development phase of mobile apps. Protocol papers and conference abstracts were excluded. Studies focusing on healthcare personnel, caregivers, or family members rather than patients were also excluded.

### Information sources

Five electronic databases were systematically searched: PubMed, Scopus, SAGE Journals, ScienceDirect, and Web of Science. In addition, reference lists of the included studies were also manually screened to identify additional eligible studies; however, none met the inclusion criteria.

### Search strategy

A comprehensive search strategy has been developed using keywords and Medical Subject Headings (MeSH) related to diabetes, chronic kidney disease, diabetic kidney disease, self-management, and mobile applications. Each database has a tailored search strategy, and Boolean operators (AND/OR) were used to refine the search queries. All databases were searched from inception to 26^th^ February 2025, with no limits on language set. An example of the whole search string is included in S2 Table.

### Study selection and data charting

All search results were imported into reference management software, Zotero, and duplicates were removed. Two reviewers have independently screened titles and abstracts against the inclusion criteria. Full-text articles of potentially relevant studies were reviewed independently. Data charting was conducted using a standardised extraction form developed by the review team, which included predefined fields based on the review objectives and PCC framework. Discrepancies between reviewers have been resolved through discussion or consultation with a third reviewer when necessary.

### Data extraction and synthesis

Relevant data were extracted into a structured form, including study characteristics, target population, mobile app features, technologies used, outcomes measures, and integration with care teams, if available.

### Data items

The following data items have been charted:

i. Author(s), year of publication, and country of study
ii. Study design and setting
iii. Target population (e.g., type 2 diabetes with chronic kidney disease)
iv. Mobile apps name
v. Core functional and details of self-management features
vi. Technology used and usage of supportive technologies (e.g., sensors or wearables)
vii. Integration with care team and
viii. Outcomes measurement used

### Quality assessment

Consistent with the scoping review methods and objective of this scoping review which are to identify and map the key features of mobile applications used for self-management among individuals with diabetic kidney disease, this study focused on providing a broad overview of the available evidence [9,10]. As per JBI guidelines, critical appraisal was not conducted, in line with the exploratory objectives of the review.

## Results

A total of 11 studies [11,12,13,14,15,16,17,18,19,20,21] met our review criteria. Fig 1 provides a flow diagram of article identification and inclusion.

**Fig 1.**
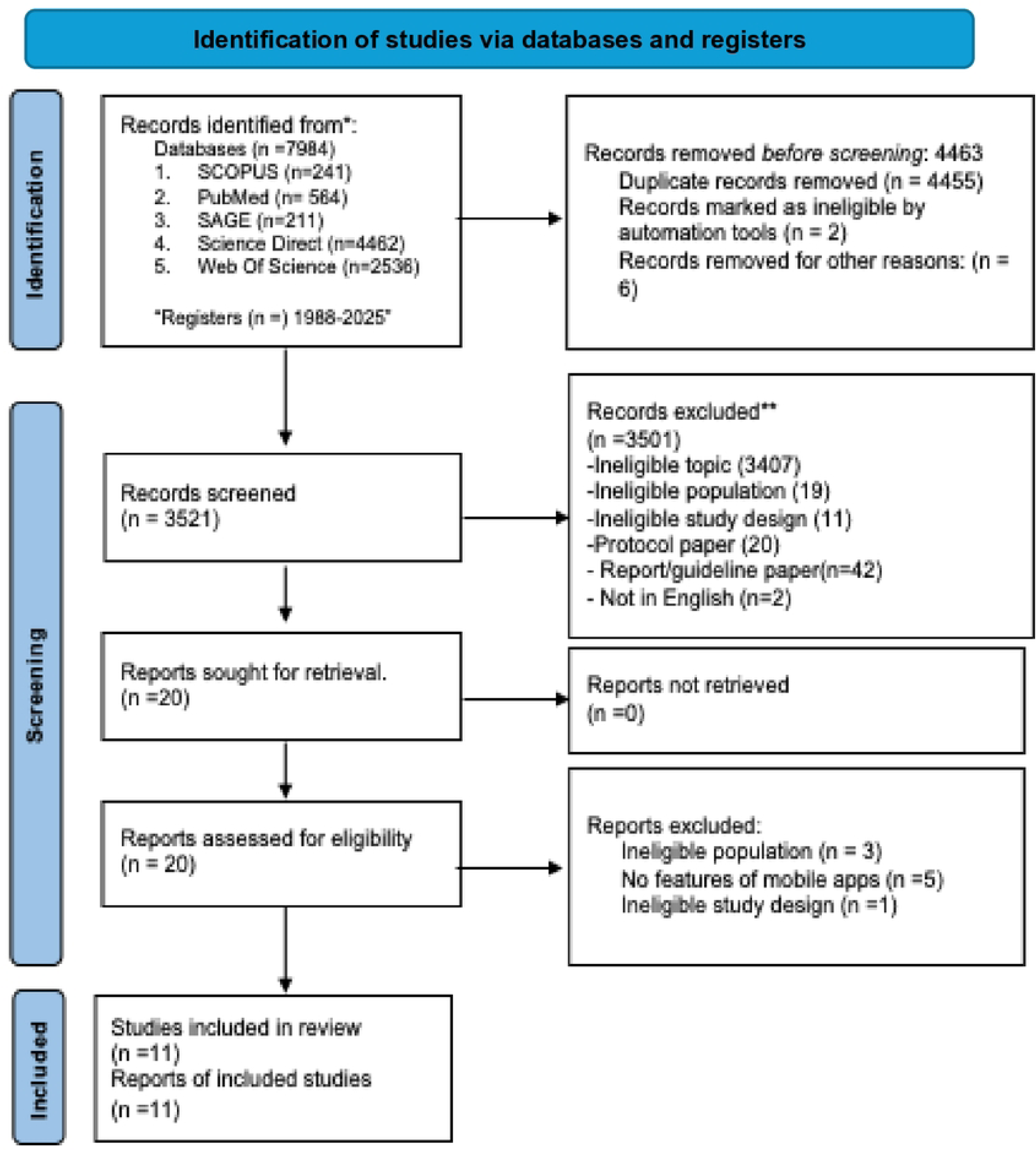
Flow diagram of article selection.

Table 2 summarises the main characteristics of the 11 studies included in this scoping review. These studies varied in terms of country of origin, year of publication, study design, target population, and the specific self-management features integrated into the mobile applications. Several studies also reported the use of supportive devices to enhance the app’s functionality. This overview provides context on the diversity of approaches employed in designing and implementing mobile health interventions for patients with diabetes, CKD, or DKD across different healthcare settings.

**Table 2.** Characteristics of included studies.

| Study No. | Article | Country | Year | Study design | Target population | Main self-management features | Other supportive devices used |
| --- | --- | --- | --- | --- | --- | --- | --- |
| 1. | Lightfoot et al. [11] | Canada | 2022 | Intervention co-development | CKD patients | i. Self-care monitoring<br>ii. Educational<br>iii. Patient's support/motivational | None reported |
| 2. | Ong et al. [12] | United States | 2021 | a randomised clinical trial | CKD patients | i. Self-care monitoring<br>ii. Patient's support/motivational | None reported |
| 3. | Li et al. [13] | Taiwan | 2020 | a randomised clinical trial | CKD patients | i. Self-care monitoring<br>ii. Patient's support/motivational | i. Heart rate wristband (detect steps, calories, sleep)<br>ii. LINE app group - to deliver diet and exercise information and teleconsultations |
| 4. | Waki et al. [14] | Japan | 2024 | a randomised clinical trial | DKD Patients | i. Self-care monitoring<br>ii. Patient's support/motivational | Bluetooth glucometer |
| 5. | Markossian et al. [15] | United States | 2021 | User-centred design-development study | CKD patients | i. Self-care monitoring<br>ii. Educational<br>iii. Patient's support/motivational<br>iv. Performance incentive | None reported |
| 6. | Toapanta et al. [16] | Spain | 2024 | Validation-Pilot monitoring study | DKD Patients | i. Self-care monitoring<br>ii. Educational<br>iii. Patient's support/motivational | chat message/video call |
| 7. | Zhang et al. [17] | China | 2024 | a randomised clinical trial | T2DM | i. Self-care monitoring<br>ii. Educational<br>iii. Patient's support/motivational<br>iv. Performance incentive | None reported |
| 8. | Hidalgo et al. [18] | Spain | 2014 | System Design & Technical Validation (Descriptive study) | T1DM<br>T2DM | i. Self-care monitoring<br>ii. Educational<br>iii. Patient's support/motivational | None reported |
| 9. | Tsai et al. [19] | Switzerland | 2021 | Prospective Cohort Study | CKD patients | i. Self-care monitoring<br>ii. Educational<br>iii. Patient's support/motivational | None reported |

| **Study No.** | **Article** | **Country** | **Year** | **Study design** | **Target population** | **Main self-management features** | **Other supportive devices used** |
| --- | --- | --- | --- | --- | --- | --- | --- |
| 10. | Berlot et al.<br>[20] | New York,<br>US. | 2024 | development and evaluation study<br>- single-arm trial study | T2DM | i. Educational<br>ii. Patient's support/motivational | None reported |
| 11. | Gunawarde<br>na et al.<br>[21] | Sri Lanka | 2019 | a randomized clinical trial | T2DM | i. Self-care monitoring<br>ii. Patient's support/motivational | Bluetooth glucometer |

### Study methods

The studies included in this review employed various research designs. Five studies utilized a randomized clinical trial (RCT) design [12,13,14,17,21] while three studies adopted user-centered or co-development approaches [11,15,20]. One study conducted a pilot validation [16] another employed a prospective cohort design [19] and one study carried out a descriptive technical validation [18].

### Year of publication and target population

The included studies were published between 2014 and 2024, with a concentration of research conducted in the past five years from 2020 to 2024. The earliest study was by Hidalgo et al. [18] in 2014, while the most recent were published in 2024. The studies targeted patients with a range of chronic conditions, primarily focusing on chronic kidney disease (CKD), diabetic kidney disease (DKD), and type 2 diabetes mellitus (T2DM). As shown in Fig 2 specifically, five studies involved CKD patients [11,12,15,19], two focused on DKD patients [14,16] and four studies targeted individuals with T2DM [17,18,20,21]. One study [18] included both T1DM and T2DM patients. This diversity in target populations reflects the overlapping and interrelated nature of these chronic conditions in mobile health research.

**Fig 2.**
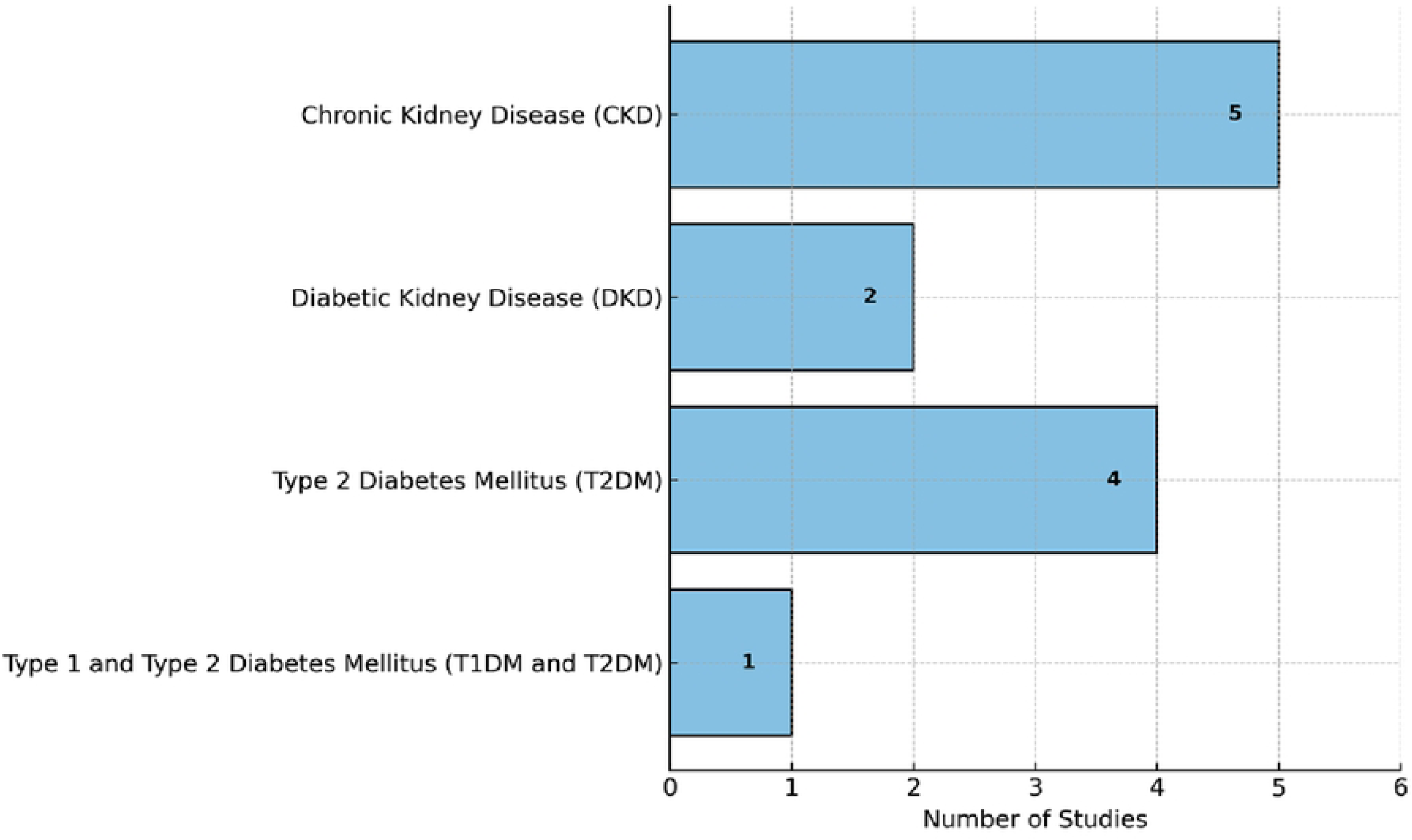
Number of studies by condition.

### Core functional domains of self-management features

Table 3 shows four recurring core functional domains of self-management features were identified across the studies. As shown in Table 3, self-care monitoring was present in ten studies, allowing users to track parameters such as blood glucose, blood pressure, and physical activity. Educational components were featured in seven studies, providing patients with disease-specific knowledge and lifestyle guidance. All eleven studies included patient support and motivational features, such as reminders, feedback, and communication tools. Lastly, performance incentive features, such as goal tracking and gamification, were incorporated in two studies [15,17] aimed at encouraging adherence and engagement.

**Table 3.**
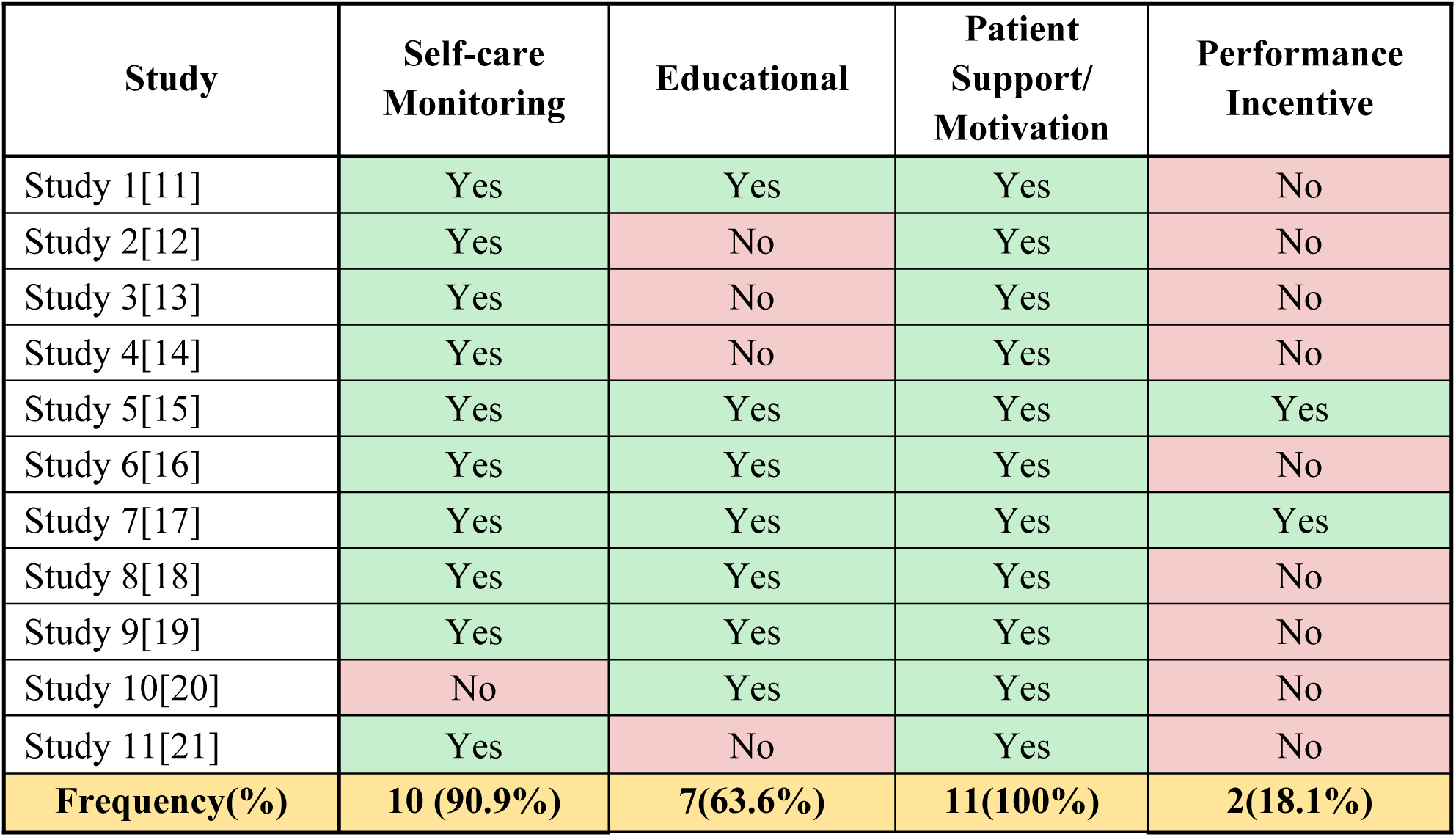
Core functional domains across the 11 studies.

Further details of the four key self-management domains identified across the included studies are presented in Table 4. These domains reflect the core functionalities integrated into mobile health interventions aimed at enhancing chronic disease self-management. Among the 11 studies, patient support and motivational features were universally implemented. These included reminders, personalized feedback, peer interactions, and clinician alerts tools known to enhance adherence and user engagement. Self-care monitoring was nearly as prevalent, appearing in 10 studies [11,12,13,14,15,16,17,18,19,21] with features such as self-monitoring blood glucose, blood pressure, pulse and activity tracking, which some supported by wearable technology.

**Table 4.** Details of core functional domains for self-management, integration, and outcome measures.

| No. | Study | Mobile apps name | Core functional domains of self-management features description |  |  |  | Integration with Care Team |  | Outcome Measures |
| --- | --- | --- | --- | --- | --- | --- | --- | --- | --- |
|  |  |  | Self-care | Educational | Patient's support/ motivational | Performance incentive | Availability | Details |  |
| 1 | Lightfoot et al. [11] | My Kidney & Me (MK&M) | i.Blood pressure<br>ii.step counting<br>iii.healthy eating<br>iv.goal setting features | -Videos:<br>i. the kidneys<br>ii. Kidney disease<br>iii. kidney disease and general health<br>iv.treatment options available<br>v. reducing my health risks<br>vi. moving more and being active<br>vii. Keeping my muscles healthy<br>viii. eating a healthy balanced diet<br>ix. managing my symptoms<br>x. Improving my sleep quality<br>xi. Looking after my well being<br>xii. Goal setting<br>-Learning session<br>-Quizzes | Tailored education, goal setting, behavior change support, patient forum | None reported | Yes | Minimal—no real-time communication or data sharing with healthcare providers. | Adherence, BP control |
| 2 | Ong et al. [12] | e-KidneyCare | i. Blood pressure<br>ii.Assessing symptoms<br>iii. Connected with pharmacy database<br>iv. Blood test: eGFR, hemoglobin, potassium, phosphate | None | Personalized feedback, lab and symptom tracking, provider alerts | None reported | Yes | Integrated alert system data from the app triggered alerts to healthcare teams when thresholds were exceeded. | HbA1c reduction, Adherence |
| 3 | Li et al. [13] | WowGoHealth app | i. Blood pressure<br>ii.heart rate<br>iii.steps counting | thru support group- none in app | Peer support via social media group, personalized | None reported | Yes | Moderate integration: data analytics provided trend summaries | Weight loss, BP, HbA1c |
|  |  |  | Self-care | Educational | Patient's support/ motivational | Performance incentive | Availability | Details |  |
|  |  |  |  |  | advice, goal tracking |  |  | for care teams feedback. |  |
| 4 | Waki et al. [14] | DialBetes Plus | i.step counts<br>ii.blood glucose<br>iii. Blood pressure<br>iv.body weight<br>v.dietary evaluation feedback | None | Self-monitoring with feedback, lifestyle guidance, in-app feedback | None reported | Yes | Data synchronized with electronic medical records (EMR) and viewable by physicians during appointments. | Glucose level, Adherence |
| 5 | Markossian et al. [15] | N/A | i. Blood pressure<br>ii.Weight<br>iii.Blood glucose<br>iv.Medication reminder | Info on<br>i. CKD<br>ii. Nutrition and recommended diets<br>iii. Concept of self-management<br>iv. Anxiety and depression<br>v. Covid-19 symptoms and resources<br>vi. Symptoms of CKD | Health tracking with feedback, real-time provider interaction, education | Gamified or performance-based incentives (non-financial) | No | No integration: app was patient-facing only, focused on goal setting and self-motivation. | Goal adherence, Weight loss |
| 6 | Toapanta et al. [16] | NORA application | i.Height/weight<br>ii. Self Monitoring Blood Glucose (SMBG)<br>iii.SpO2<br>iv. Blood pressure<br>v. steps count<br>vi. Medication reminder | Health education material:<br>i. Education module-managing blood pressure, blood glucose and weight.<br>Medication adherence<br>ii. FAQ | Mood check-ins, clinician messaging, educational info, adherence support | None reported | Planned for future | No integration—developmental phase, care team involvement not yet implemented. | App usability |
|  |  |  | Self-care | Educational | Patient's support/ motivational | Performance incentive | Availability | Details |  |
| 7. | Zhang et al. [17] | SMARTDiabetes | i. Blood pressure<br>ii.BMI<br>iii. Medication reminder<br>iv. Blood test: Glucose, LDL, Cholesterol, HbA1c | The information are organized into seven sections:<br>I. General noncommunicable diseases,<br>ii.Diabetes,<br>iii. Hypertension,<br>iv.Weight control,<br>v.Physical exercise,<br>vi.Other lifestyle<br>vii.Physical examination. | Personalized goals, reminders, family support, education | Small incentives for participation (gift cards, social recognition) | Yes | Real-time data synced to central dashboard accessible by care team to monitor patient progress. | BP,HbA1c, App adherence |
| 8. | Hidalgo et al. [18] | glUCModel | i.diet<br>ii.sport activity (scale level of exercise 1-10 + introduce the duration of the exercise and complete a remark field explaining the sport activity performed.)<br>iii.weight<br>iv.BMI<br>v. Insulin reminder<br>vi. Blood test- glucose level | Information on<br>i.Hypoglycaemia management,<br>ii. Diet,<br>iii. Symptoms | Personalized recommendations, education, feedback engine | None reported | Planned for future | Planned for future—currently no integration but designed with provider-facing potential. | Usability, Acceptability |
|  |  |  | Self-care | Educational | Patient's support/ motivational | Performance incentive | Availability | Details |  |
| 9. | Tsai et al. [19] | iCKD | i. Blood pressure<br>ii. weight<br>iii. exercise<br>iv. meal photo<br>v. Medication compliance<br>vi. Blood test: SMBG, Blood urea nitrogen (mg/dL) eGFR (ml/min/1.73 m<sup>2</sup>) Hemoglobin (g/dL) Albumin (g/dL) Uric acid (mg/dL) Cholesterol (mg/dL) Triglyceride (mg/dL) Urine protein/creatinine ratio (mg/mg) HbA1c (%) | Educational video<br>i. Kidney anatomy & physiological functions,<br>ii. Symptoms of CKD, and<br>iii. diagnostic indicators such as blood pressure and laboratory values (e.g., eGFR, hemoglobin) | Education modules, self-care feedback, reminder alerts | None reported | Yes | Moderate—tracking dashboard accessible by clinic staff to review motivation and biometric data. | A1c, Motivation scores |
| 10. | Berlot et al. [20] | Diabetes Xcel mobile app | none | Animated videos + modules<br>1 “What are the different types of diabetes and who is at risk?”<br>2 “Hyperglycemia and diabetes-related problems”<br>3 “How can I manage my diabetes with the ABCs?”<br>4 “How can I plan what to eat?”<br>5 “Diabetes and exercise”<br>6 “How can I monitor my diabetes control?”<br>7 “Diabetes medications”<br>8 “Tests your health care provider will do”<br>9 “Resources” | Supportive push messages, video-based coaching, reminders | None reported | Yes | Minimal—educational prompts without two-way provider communication. | Adherence, Knowledge gain |
|  |  |  | Self-care | Educational | Patient's support/ motivational | Performance incentive | Availability | Details |  |
| 11. | Gunawardena et al. [21] | Smart Glucose Manager | i. Diet reminder<br>ii. Exercise at user-defined times<br>iii. Medication reminders<br>iv. SMBG | None | Automated reminders, bolus insulin calculator, self-care tracker | None reported | Yes | High integration—telemedicine-based model with real-time counseling chat and provider alerts. | Glucose control, Patient satisfaction |

Educational content was incorporated in seven studies [11,15,16,17,18,19,20], delivered through interactive modules, videos, or structured text, supporting patient understanding of disease processes and lifestyle management. In contrast, performance incentives such as gamification or reward systems were reported in only two studies which one was conducted in United States [15] and the other in China [17].

### Supportive devices used

In addition to the use of basic mobile applications, a limited number of studies incorporated other supportive devices to enhance the functionality and user experience of self-management interventions. Out of the eleven studies reviewed, four reported the integration of additional tools. One study utilized a heart rate wristband to monitor physical activity parameters such as steps taken, calories burned, and sleep patterns [15]. Two studies employed the LINE app group function to deliver dietary and exercise information and to facilitate teleconsultations [13,16]. Another two studies incorporated Bluetooth-enabled glucometers, allowing users to synchronize blood glucose data directly with the mobile application for continuous monitoring [14,21]. Study 6 [16] integrated communication features, including chat messaging and video calls, to provide real-time interaction and support. The remaining studies did not report the use of external supportive devices and relied solely on the mobile application’s built-in features.

### Integration with care team and outcome measures

Among all the 11 studies, only a minority demonstrated active integration between the mobile apps and healthcare providers. Some apps (e.g., DialBetesPlus and SMARTDiabetes) synchronized data with electronic medical records or allowed care teams to monitor patient progress [14,17]. Others (e.g., e-KidneyCare) used built-in alerts to notify providers of critical lab results or symptoms [12]. However, most apps were standalone tools, lacking bidirectional communication or real-time clinician input. The included studies reported a range of outcome measures encompassing adherence metrics (such as medication compliance and goal tracking), biometric improvements (including blood pressure, HbA1c, and weight), and patient-reported outcomes related to usability and satisfaction [11,21]

## Discussion

This scoping review aimed to identify and characterize the features of mobile applications designed to support self-management among individuals with diabetic kidney disease (DKD), type 2 diabetes mellitus (T2DM), and chronic kidney disease (CKD). Given the pathophysiological overlap between these conditions particularly DKD as a progressive outcome of both T2DM and CKD, mobile apps developed for any of these populations often address similar self-management challenges. Across the reviewed studies, the most consistently implemented features included patient support and motivational tools, self-care monitoring functions, and educational components.

Patient support mechanisms such as reminders, personalised feedback, peer interaction features, and messaging were universally implemented across all included studies. These features are central to enhancing treatment adherence, reinforcing lifestyle behaviours, and sustaining long-term engagement. Their consistent inclusion reflects a paradigm shift toward interactive, behaviorally informed, and patient-centred app design. This trend aligns with a growing body of evidence suggesting that supportive digital communication significantly improves adherence and health outcomes in diabetes and kidney care [1,22].

Self-care monitoring was also a highly prevalent domain, reported in 10 of the 11 studies. These functionalities enabled users to track various health parameters such as blood glucose, blood pressure, body weight, and physical activity either manually or through wearable device integration. Some apps, for example, employed Bluetooth-enabled glucometers, enhancing real-time data capture and user convenience. These features support the established role of physiological monitoring in optimizing glycemic and cardiovascular outcomes for patients with diabetic complications [2].

Educational content was featured in seven studies, often delivered via structured videos, app-based modules, or interactive quizzes. These components aimed to improve disease understanding, medication adherence, dietary practices, and symptom recognition. However, the degree of personalization and interactivity varied substantially. This highlights a current gap in the use of dynamic, adaptive learning systems that tailor educational content to users’ evolving needs as an area deserving further innovation and research [23,24].

A notable but underutilized design element identified in this review is the inclusion of performance incentives, such as gamification and reward mechanisms. These were reported in only two studies, despite emerging research demonstrating their potential to enhance behavioral outcomes and sustained user engagement [3]. Gamification elements especially achievement tracking and competition can increase motivation but must be balanced to avoid user fatigue and app abandonment [25].

A recurring trend observed in the studies was the minimal emphasis on enabling real-time communication between patients and healthcare providers. Although some apps included features like data sharing or provider alerts, very few enabled two-way, live interactions. This is a missed opportunity, especially as healthcare increasingly moves toward more connected, team-based care. Digital tools that support continuous, bidirectional communication are essential not only for urgent issues, but also to sustain long-term chronic disease management. Integrated digital systems that promote seamless provider-patient interaction are crucial for minimizing care fragmentation and improving chronic care outcomes, as highlighted by recent evaluations of digital health strategies [26]. The significance of culturally tailored integrative application design incorporating personalized education and real-time communication is demonstrated by the mobile app developed for patients with type 2 diabetes mellitus in Indonesia [27]. These examples underscore the value of embedding clinical integration and interactive support within mobile health solutions. Looking ahead, app developers should aim to blend self-management features with seamless communication and personalized support to truly enhance care continuity and patient experience.

A variation in mobile app features appeared to align with the target population and geographic region. Mobile apps developed for diabetes populations (e.g., SMARTDiabetes in China [17], glUCModel in Spain [18], and Diabetes Xcel in the United States [20]) placed greater emphasis on biometric monitoring and educational content related to glycemic control and lifestyle. In contrast, CKD mobile apps like My Kidney & Me (Canada) [11] and iCKD (Switzerland) [19] focused more heavily on symptom tracking, renal-specific lab monitoring, and general wellness modules. Meanwhile, mobile apps targeting DKD (DialBetesPlus in Japan [14] and NORA in Spain [16]) incorporated hybrid functionalities that reflect the complex self-management needs of individuals living with both diabetes and kidney disease. These apps combined monitoring tools for blood glucose, blood pressure, body weight, and oxygen saturation (SpO₂), thus addressing both glycemic control and renal health. Regionally, Asian-developed apps (e.g., WowGoHealth in Taiwan [13] and Smart Glucose Manager in Sri Lanka [21]) tended to include wearable device integration and messaging features, possibly reflecting greater comfort with technology use and healthcare digitization in those contexts. This diversity implies that both the health priorities of target populations and regional technological infrastructure may shape how mobile app functionalities are prioritized.

Although outcomes such as adherence, glycemic control, or usability were reported in several studies, this review did not evaluate their effectiveness. Instead, our focus remained on mapping the functional features and structural design elements of mobile apps to support self-management in DKD, T2DM, and CKD populations. This approach aligns with other recent reviews that prioritize descriptive analysis of app content and functionality over impact assessment, particularly in fields where intervention trials remain limited or heterogeneously reported [28].

Like any scoping review, this study has some limitations that are important to acknowledge. First, we did not perform a formal quality assessment of the included studies. This is consistent with the Joanna Briggs Institute’s scoping review methodology, which focuses on mapping existing evidence rather than evaluating study rigor. However, it means that the methodological quality of the reviewed studies may vary, and our findings should be interpreted with that in mind. Second, the review included only studies published in English and indexed in selected databases. This may have led us to miss relevant mobile applications described in non-English literature or grey literature such as app store listings or unpublished reports. Another key point is that our review centered on describing the features and design elements of mobile apps, not their clinical effectiveness. Despite these limitations, our study is among the first to comprehensively explore mobile health apps spanning DKD, T2DM, and CKD. By comparing self-management features across these overlapping conditions, this review highlights underutilized functionalities such as gamification and clinical integration and offers practical guidance for designing more connected, personalized, and engaging digital health tools.

## Conclusion

This scoping review mapped the core self-management features of mobile applications designed for DKD, DM and CKD populations. While patient support tools, self-monitoring capabilities, and educational components were widely implemented, few apps integrated motivational incentives or direct communication with healthcare providers. These findings highlight the importance of incorporating bi-directional data sharing between users and healthcare providers to support real-time clinical oversight and personalized feedback. Additionally, the inclusion of behavioral reinforcement strategies such as adaptive goal setting and incentive-based features can enhance user motivation and long-term engagement. Insights from this review may serve as a valuable resource for digital health developers, nephrology and diabetes care teams, and health policymakers aiming to design more patient-centered, functionally integrated mobile health interventions.

## Data Availability

All relevant data are within the manuscript and its Supporting Information files.

## Acknowledgement

This work was supported by the Universiti Sains Malaysia, Research University Team (RUTeam) Grant Scheme with Project No: 1001/PPSP/8580083, Project Code: TE0034 (Reference No: 2022/0501]

## Author contributions

**Project administration & Resources:** Nani Draman

**Data curation:** Hasmawati Yahya, Nani Draman, Najib Majdi Yaacob.

**Methodology:** Hasmawati Yahya, Azidah Abdul Kadir, Najib Majdi Yaacob.

**Writing – original draft:** Hasmawati Yahya

**Writing, review and editing:** Hasmawati Yahya, Azidah Abdul Kadir. Najib Majdi Yaacob.

## Supporting information

**S1 Table.**
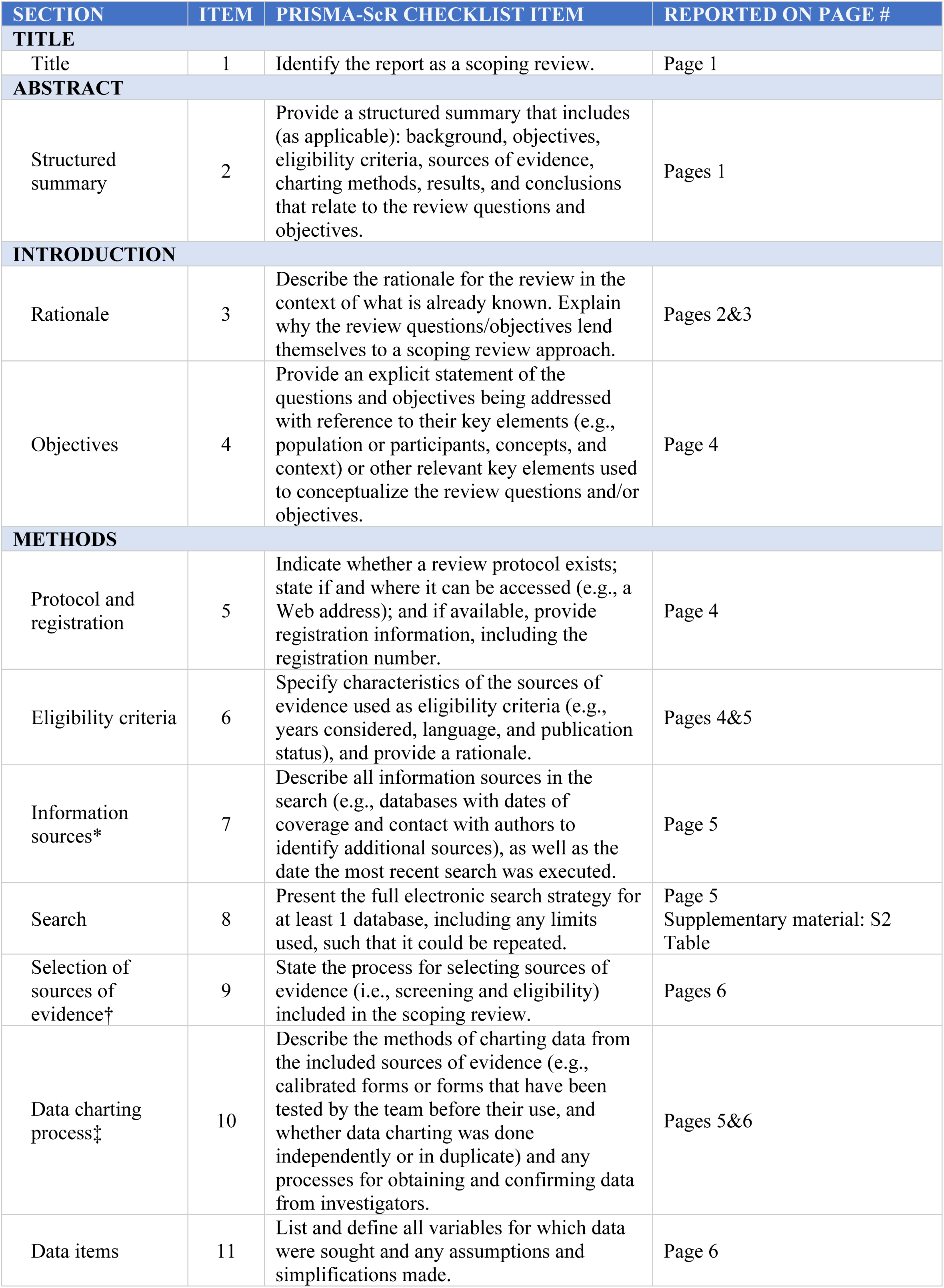
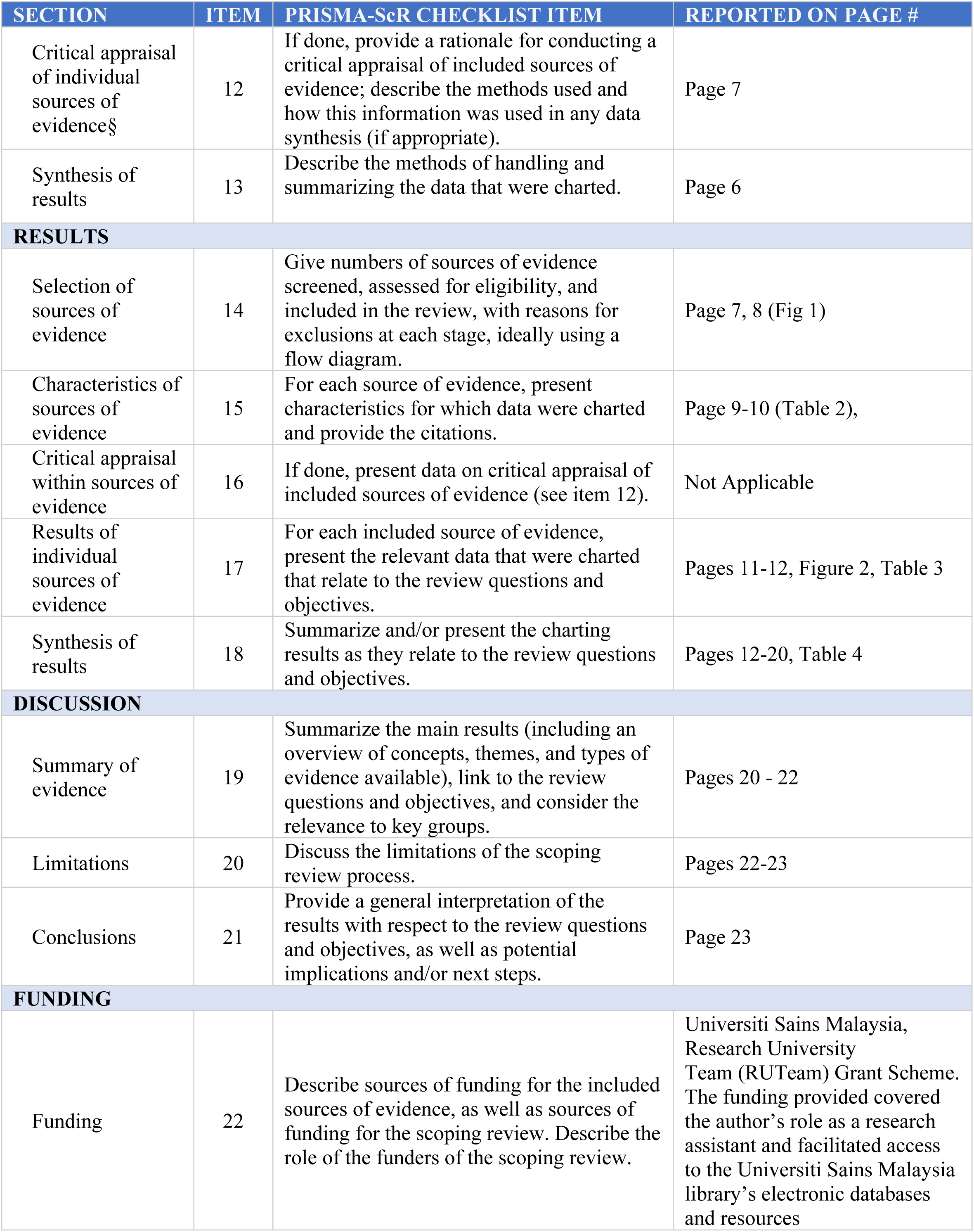
Preferred reporting items for systematic reviews and meta-analyses extension for scoping reviews (PRISMA-ScR) checklist.

**S2 Table.**
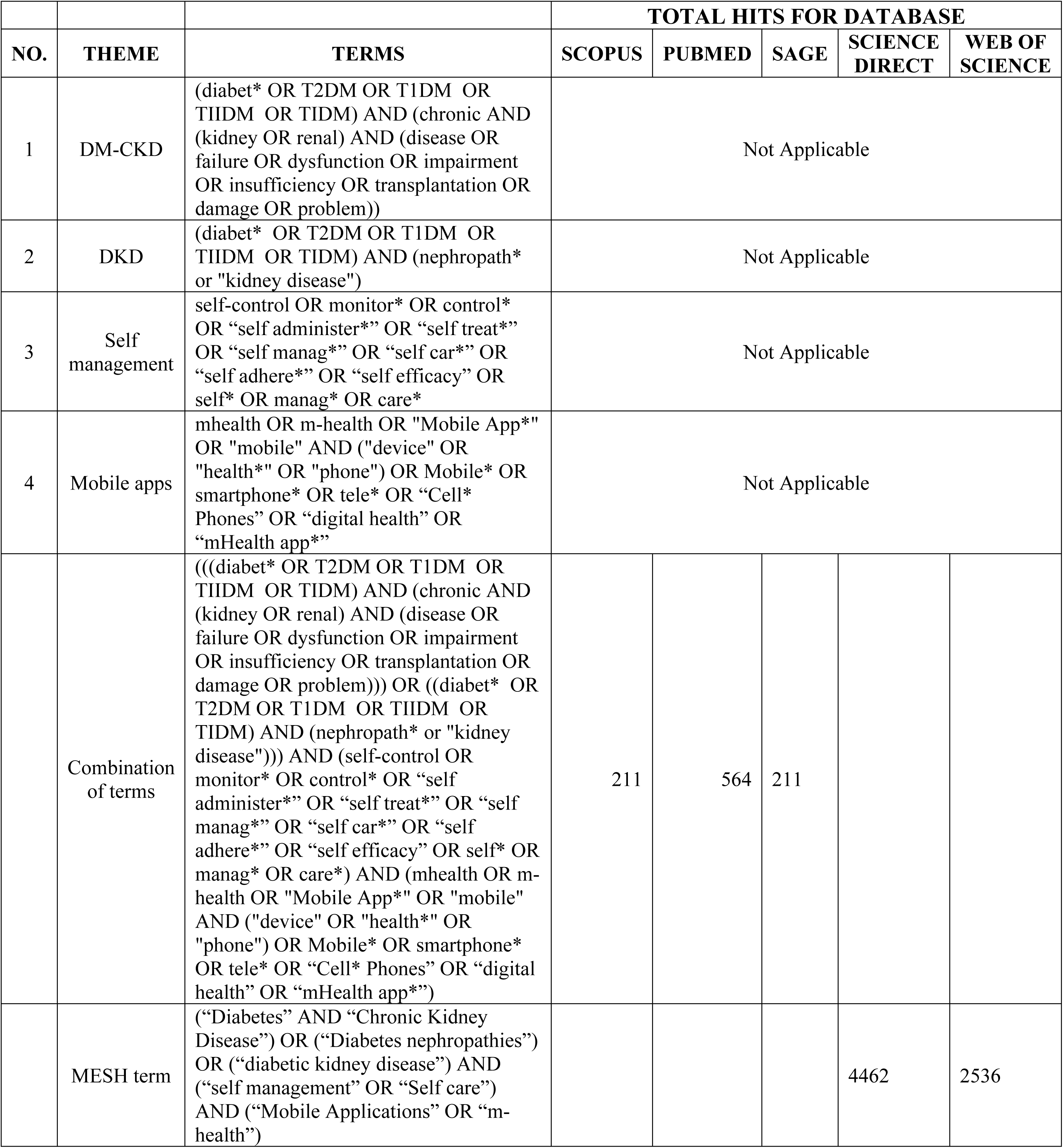

